# Comparative Efficacy and Safety of Coronary Artery Bypass Grafts: Insights from a Systematic Review and Meta-Analysis

**DOI:** 10.1101/2025.09.20.25336224

**Authors:** Ritesh Mate, Aftab Hossain, Rakhshanda Khan, Rohith Kumar, Krithika Nathan, Y. Vedhanth Reddy, Kavya Lnu, Harshaardhan Dhanraj Ramteke

## Abstract

**Introduction:** Optimal conduit selection in coronary artery bypass grafting (CABG) remains debated. While the left internal mammary artery (LIMA) is standard, the comparative performance of bilateral internal mammary arteries (BIMA), radial artery (RA), right internal mammary artery (RIMA), and saphenous vein grafts (SVGs) has not been comprehensively established. We performed a meta-analysis of observational studies to evaluate the efficacy, safety, and outcomes of different coronary grafts.

**Methods:** A systematic search identified **795 studies**, of which **32 observational studies** comprising **172,911 patients** were included. Data were pooled using random-effects models in STATA. Outcomes included survival, graft occlusion, graft failure, patency, revascularization, mortality, adverse events, wound infection, length of stay, and renal function.

**Results:** A total of **32 observational studies** involving **172,911 patients** (69,402 male and 100,658 female; mean follow-up 105.1 months) were included. The most frequently used conduit was the **LIMA (66,135 patients)**, followed by **SVG (48,892)**, **BIMA (20,133)**, **RA (8,516)**, **RIMA (2,305)**, **LIMA+SVG (2,339)**, and **LIMA+RA (60)**. BIMA grafting was associated with significantly improved survival (log OR –0.68; 95% CI –0.93 to –0.44) and reduced graft failure (–0.67; 95% CI –1.02 to –0.32), whereas SVGs demonstrated worse survival (–3.94; 95% CI –4.07 to –3.82) and lower patency (–4.00; 95% CI –4.44 to –3.56). RA grafts reduced cardiac mortality (–1.30; 95% CI –1.89 to –0.71) and adverse events (–0.69; 95% CI –1.09 to – 0.28), while RIMA was associated with increased graft failure (2.50; 95% CI 1.47–3.52) and higher all-cause mortality (1.09; 95% CI 0.60–1.58). SVGs lowered the risk of sternal wound infections (–1.77; 95% CI –2.61 to –0.92), while BIMA showed a nonsignificant increase. BIMA also shortened hospital stay (–0.83 days; 95% CI –1.45 to –0.20). No significant differences were observed across conduits for renal dysfunction.

**Conclusion:** This meta-analysis confirms the superiority of arterial grafts, particularly BIMA and RA, in enhancing survival and reducing adverse outcomes compared with SVGs. LIMA remains the cornerstone graft, while the use of multiple arterial grafting strategies provides durable long-term benefits. Careful patient selection is essential, particularly given the higher wound complication risk with BIMA and variable outcomes with RIMA. These findings reinforce the growing role of total arterial revascularization, though further randomized trials are warranted to refine graft selection strategies.

## Introduction

Coronary artery disease (CAD) remains the leading cause of morbidity and mortality worldwide, responsible for nearly one-third of global deaths annually [1]. Coronary artery bypass grafting (CABG) is the most established surgical intervention for patients with multivessel or left main CAD, providing superior long-term outcomes compared with percutaneous coronary intervention (PCI) in appropriately selected populations [2,3]. Despite advances in PCI and drug-eluting stent technology, CABG continues to offer improved survival and reduced major adverse cardiovascular events (MACE) in complex disease. A key determinant of CABG success is the choice of conduit for myocardial revascularization.

The left internal mammary artery (LIMA) has consistently demonstrated excellent long-term patency when grafted to the left anterior descending (LAD) artery, with landmark studies showing significant survival benefit and reduced cardiac events compared with saphenous vein grafts (SVGs) [4–6]. As a result, the LIMA is universally accepted as the gold standard graft in CABG. However, beyond the first arterial conduit, the optimal choice of additional grafts remains debated. SVGs are still the most commonly used conduits worldwide due to their technical simplicity and ready availability. Yet, their durability is limited: up to 50% of SVGs occlude within ten years, raising concerns about their long-term efficacy [7]. In contrast, other arterial grafts, such as the right internal mammary artery (RIMA), the radial artery (RA), and the gastroepiploic artery, have shown superior patency and potential survival advantages [8–10]. The use of bilateral internal thoracic arteries (BITA) has been advocated to improve long-term outcomes, but widespread adoption has been hampered by increased surgical complexity and higher risks of sternal wound infection, particularly among diabetics and obese patients [11,12].

Over the past two decades, several systematic reviews and meta-analyses have attempted to clarify the relative benefits of different grafts. Some Studies reported improved outcomes with arterial conduits, especially radial arteries, compared with SVGs [13,14]. A study [15] demonstrated that RA grafting conferred better patency and survival than venous grafts, particularly in patients with high-grade coronary stenosis. Similarly, Another study [16] found that BITA grafting was associated with a survival benefit compared with single internal thoracic artery (SITA) grafting, although at the cost of increased wound complications. Conversely, the large randomized Arterial Revascularization Trial (ART) failed to show a significant survival difference between BITA and SITA strategies at ten years [17]. Other reviews and pooled analyses have provided conflicting evidence: some highlighted long-term survival advantages of multiple arterial grafts [18], while others emphasized that these benefits diminish when perioperative risks and patient comorbidities are taken into account [19].

The existing evidence base, while extensive, suffers from several important limitations. Most randomized controlled trials (RCTs) are underpowered to detect long-term survival differences due to small sample sizes and the lengthy follow-up required. Observational studies, though larger, are subject to confounding and selection bias. Prior meta-analyses have typically restricted their focus to specific comparisons, such as RA versus SVG or BITA versus SITA, rather than comprehensively addressing all graft options in CABG. Moreover, marked heterogeneity exists across studies regarding surgical technique (on-pump versus off-pump), patient characteristics (for example, diabetic versus non-diabetic populations), and outcome definitions. These differences complicate the synthesis and interpretation of results. Angiographic follow-up for graft patency is inconsistently reported, further limiting the strength of available evidence.

Given these uncertainties, a significant gap persists in the literature. Although multiple trials and reviews have assessed selected conduits, there is no comprehensive synthesis that evaluates all major coronary grafts within a unified analytical framework. Previous studies have emphasized survival or patency in isolation, often overlooking the balance between efficacy and safety, such as perioperative complications, wound infections, or the need for repeat revascularization. Consequently, clinical decision-making regarding optimal graft choice remains variable, influenced as much by surgical preference and institutional practice as by robust evidence.

To address this gap, the present meta-analysis aims to systematically review and synthesize the available randomized and observational data comparing all commonly used conduits in CABG. By pooling survival, patency, and safety outcomes across a wide range of studies, this analysis seeks to provide precise and clinically relevant estimates of the relative performance of different grafts. Such evidence has the potential to guide surgeons in selecting the most effective conduits, inform international guideline recommendations, and ultimately improve long-term outcomes for patients undergoing CABG.

## Methods

### Literature Search

A comprehensive literature search was conducted across PubMed, Embase, Cochrane Central Register of Controlled Trials (CENTRAL), and Web of Science from inception to September 2025. The search combined Medical Subject Headings (MeSH) and free-text terms related to “coronary artery bypass grafting,” “CABG,” “graft,” “internal mammary artery,” “radial artery,” “saphenous vein,” and “conduit.” Reference lists of relevant articles and previous systematic reviews were screened to identify additional studies. No language restrictions were applied. Both randomized controlled trials (RCTs) and observational studies reporting clinical outcomes of different coronary conduits were eligible for inclusion.

### Study Selection and Screening

All studies identified through the search were imported into reference management software, and duplicates were removed. Two reviewers independently screened titles and abstracts to exclude irrelevant articles. Full texts of potentially eligible studies were retrieved and assessed against predefined inclusion and exclusion criteria. Eligible studies included observational studies that compared outcomes of different coronary artery bypass graft conduits and reported clinical or angiographic endpoints. Discrepancies between reviewers were resolved through discussion, and when necessary, by consultation with a third reviewer. A Preferred Reporting Items for Systematic Reviews and Meta-Analyses (PRISMA) flow diagram was used to illustrate the process of study selection and screening [20]. The Protocol is registered on Prospero with the number CRD420251151159.

### Statistical Analysis

All statistical analyses were performed using Stata (version 18.0; StataCorp, College Station, TX, USA). Pooled effect estimates were calculated using either fixed-effects (Mantel–Haenszel) or random-effects (DerSimonian–Laird) models, depending on the presence of heterogeneity. Risk ratios (RRs) with 95% confidence intervals (CIs) were reported for dichotomous outcomes, while mean differences (MDs) or standardized mean differences (SMDs) with 95% CIs were used for continuous outcomes. Heterogeneity across studies was assessed using the χ² test and quantified with the I² statistic. Publication bias was evaluated using funnel plots and Egger’s regression test where ≥10 studies were available. Sensitivity analyses were conducted by excluding high-risk or small studies to test robustness of results.

### Risk of Bias Assessment

The methodological quality of all included observational studies was assessed using the Risk Of Bias In Non-randomized Studies of Interventions (ROBINS-I) tool [21]. This framework evaluates seven domains: bias due to confounding, selection of participants, classification of interventions, deviations from intended interventions, missing data, measurement of outcomes, and selection of reported results. Two reviewers independently conducted the assessment, with discrepancies resolved through consensus or by consultation with a third reviewer. Studies were categorized as having low, moderate, serious, or critical risk of bias across each domain.

### Certainty of Evidence (GRADE)

The certainty of evidence for each outcome was assessed using the Grading of Recommendations, Assessment, Development and Evaluation (GRADE) approach. As all included studies were observational, the certainty of evidence started at low and could be downgraded further for limitations in risk of bias, inconsistency, indirectness, imprecision, or publication bias. Where appropriate, the certainty could be upgraded for factors such as large effect sizes, dose-response relationships, or when all plausible confounding would reduce an apparent effect. The overall certainty of evidence was graded as high, moderate, low, or very low. A Summary of Findings (SoF) table was prepared to present the main outcomes and corresponding GRADE ratings.

## Results

A total of 795 studies were identified in the initial search, of which 32 observational studies met the eligibility criteria and were included in the present meta-analysis, encompassing 172,911 patients [22–53]. The overall study population comprised 69,402 males and 100,658 females, with a mean follow-up duration of 105.1 months. Regarding conduit distribution, the most frequently reported graft was the left internal mammary artery (LIMA), used in 66,135 patients, followed by the saphenous vein graft (SVG) in 48,892 patients, bilateral internal mammary arteries (BIMA) in 20,133 patients, and the radial artery (RA) in 8,516 patients. Less frequently reported conduits included the right internal mammary artery (RIMA) (2,305 patients), LIMA combined with SVG (2,339 patients), and LIMA combined with RA (60 patients). These distributions reflect the predominant role of arterial conduits, particularly the LIMA, in contemporary CABG, while also highlighting the continued widespread use of SVGs. Figure 1 and Table S1.

**Figure 1.**
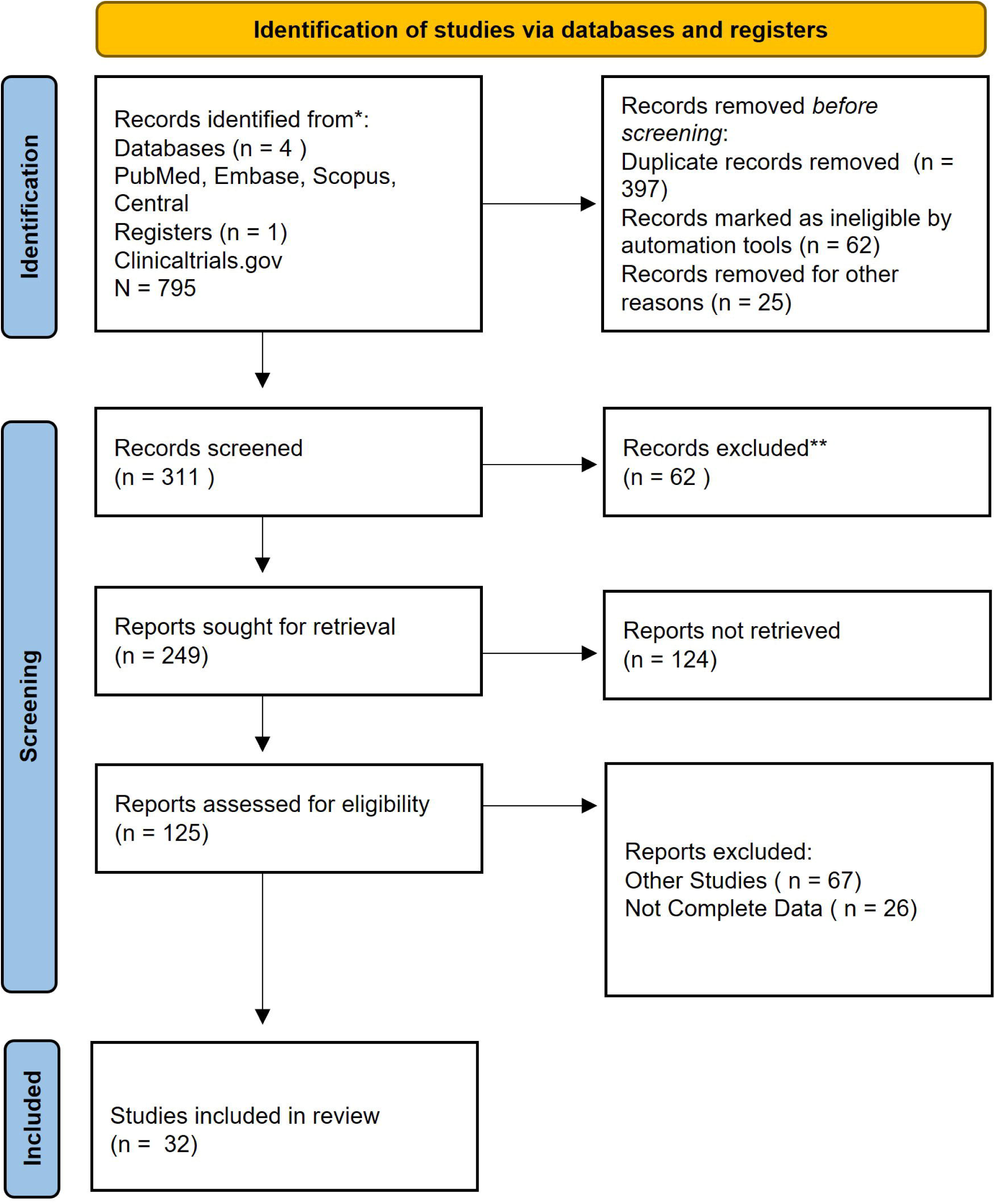
PRISMA Flow Diagram.

### Survival Outcomes According to Graft Type

The pooled analysis of 32 observational studies encompassing 172,911 patients demonstrated important differences in survival according to the conduit used (Figure 2). Bilateral internal mammary artery (BIMA) grafting was associated with a statistically significant survival advantage compared with control groups, with a pooled log odds ratio of 0.90 (95% CI, 0.34–1.46; p = 0.00), although marked heterogeneity was observed across studies (I² = 97.8%). In contrast, left internal mammary artery (LIMA) grafting alone was associated with significantly lower survival, with a pooled log odds ratio of –1.14 (95% CI, –1.63 to –0.66; p < 0.001). The combination of LIMA with saphenous vein grafts (SVGs) yielded no significant difference in survival compared with control (–0.18; 95% CI, –1.23 to 0.86). Analyses of other arterial conduits demonstrated non-significant findings. Radial artery (RA) grafts showed a directionally favorable effect (0.46; 95% CI, –0.42 to 1.33), while right internal mammary artery (RIMA) grafts also did not reach statistical significance (0.43; 95% CI, –0.71 to 1.57). By contrast, SVGs were consistently associated with markedly worse survival outcomes, with a pooled log odds ratio of –3.94 (95% CI, –4.07 to –3.82; p < 0.001). When all conduits were pooled together, the overall estimate suggested no statistically significant survival benefit (–0.06; 95% CI, –0.59 to 0.46), with substantial heterogeneity across comparisons (I² = 99.0%).

**Figure 2.**
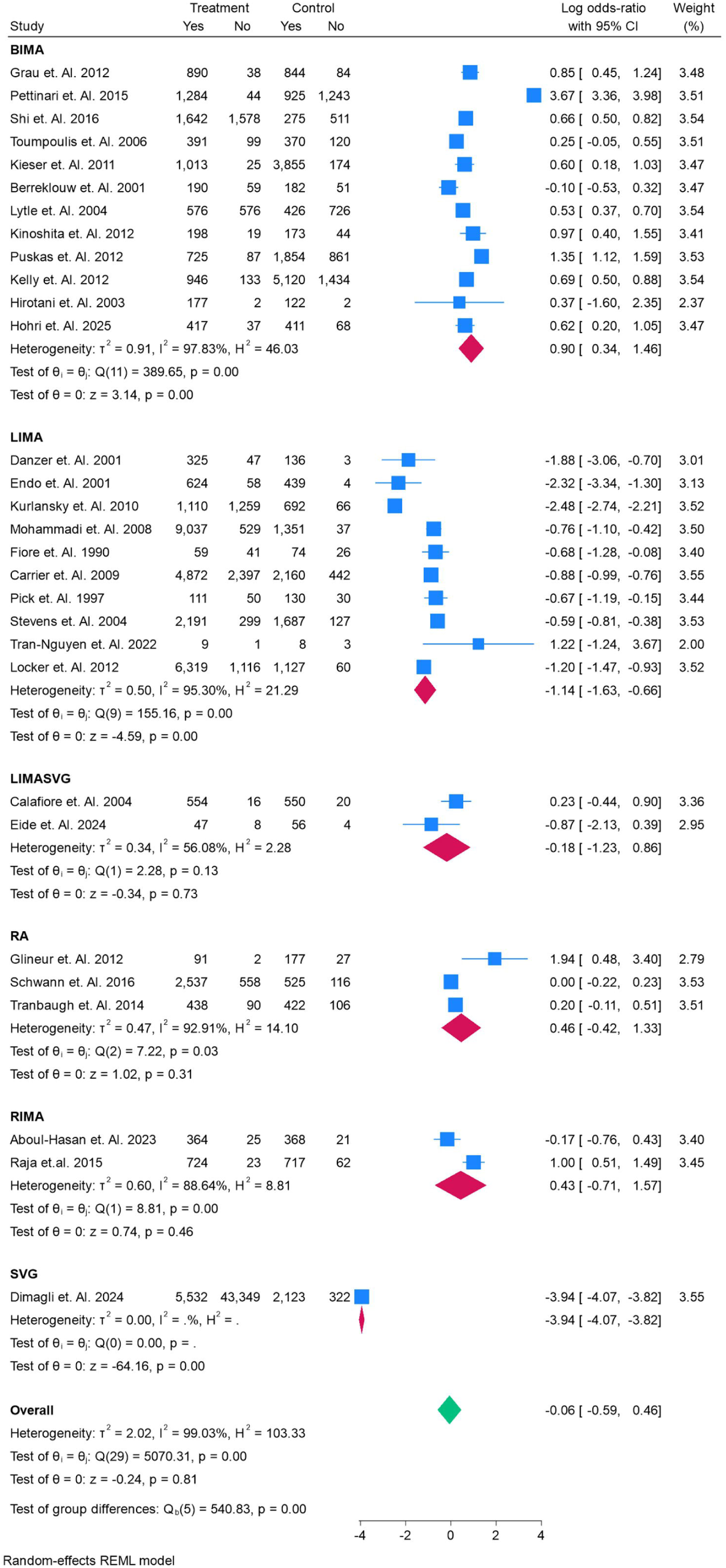
The odds of Survival of each coronary Graft in the CABG.

### Graft Occlusion Outcomes

Pooled analyses of graft occlusion demonstrated no significant overall difference between conduits (Figure 3). For bilateral internal mammary artery (BIMA) grafting, the summary log odds ratio was –0.23 (95% CI, –1.08 to 0.62; p = 0.59), with substantial heterogeneity across included studies (I² = 96.1%). Left internal mammary artery (LIMA) grafts also showed no significant association with occlusion, with a pooled estimate of 0.19 (95% CI, –1.22 to 1.60; p = 0.79). The combination of LIMA with saphenous vein grafts (SVGs) yielded a non-significant reduction in occlusion risk (–0.62; 95% CI, –3.05 to 1.81). Radial artery (RA) grafts demonstrated a neutral effect (0.08; 95% CI, –0.26 to 0.43), while right internal mammary artery (RIMA) grafts also showed no significant difference (0.39; 95% CI, –0.11 to 0.89). The overall pooled estimate across all conduit types was 0.05 (95% CI, –0.44 to 0.53; p = 0.86), again with high heterogeneity (I² = 95.7%). Thus, while individual studies suggested potential differences, meta-analytic evidence did not reveal a consistent survival advantage of any single conduit type with respect to graft occlusion.

**Figure 3.**
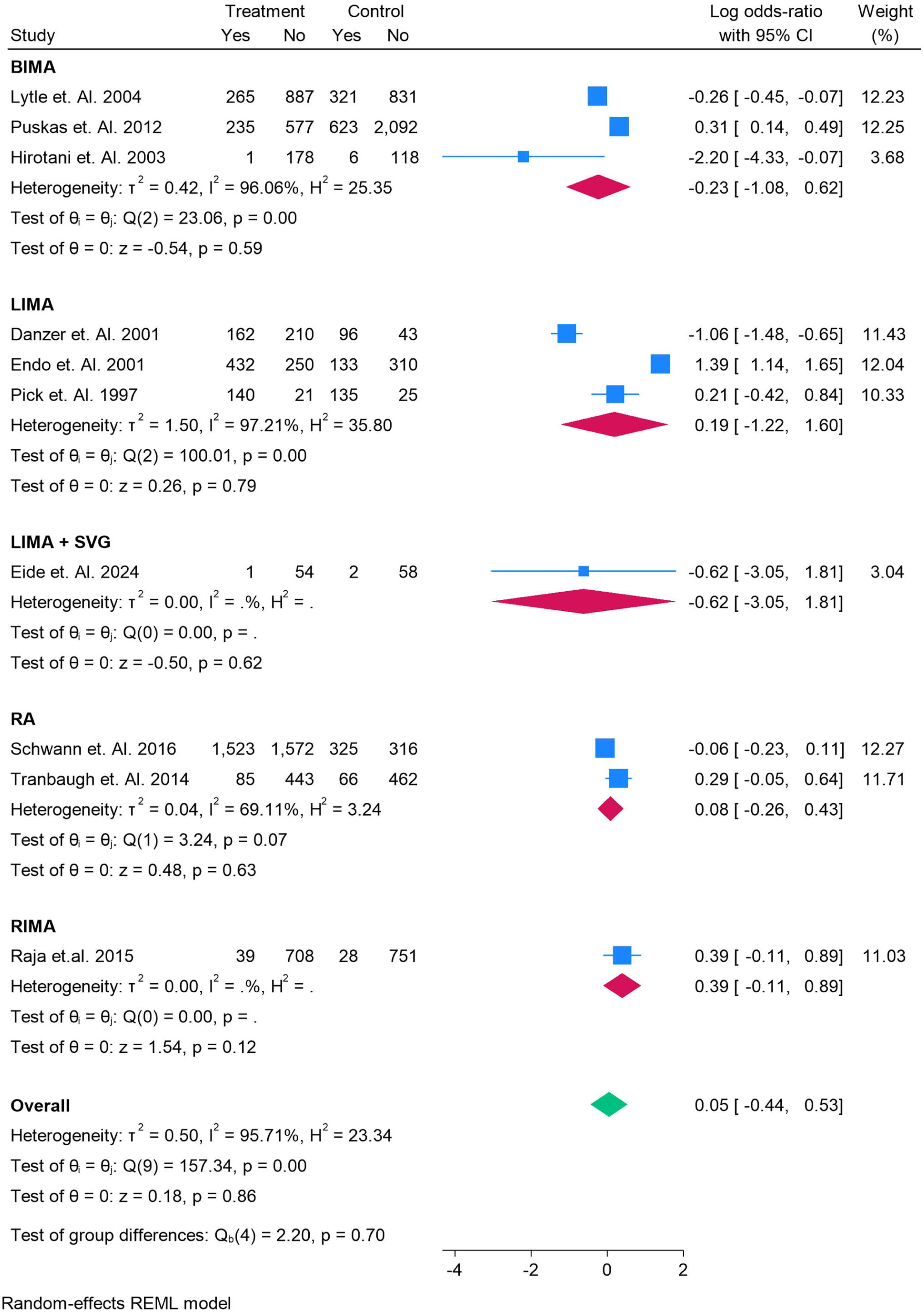
The odds of Occlusion of each coronary Graft in the CABG.

### Graft Failure Outcomes

Analysis of graft failure demonstrated marked variability across conduit types (Figure 4). Bilateral internal mammary artery (BIMA) grafts were consistently associated with a reduced risk of failure, with a pooled log odds ratio of –0.67 (95% CI, –1.02 to –0.32; p < 0.001), and no evidence of heterogeneity (I² = 0%). Similarly, LIMA grafts showed a significant association with increased graft durability, though in the opposite direction: pooled estimates indicated a higher risk of failure (0.76; 95% CI, 0.35 to 1.17; p = 0.00), suggesting variability in performance depending on study context. The combination of LIMA with SVGs showed no significant association with graft failure (–1.04; 95% CI, –3.34 to 1.25; p = 0.37). Radial artery (RA) grafts demonstrated a non-significant protective trend (0.29; 95% CI, –0.05 to 0.64), whereas RIMA grafts were significantly associated with an increased risk of failure (2.50; 95% CI, 1.47 to 3.52; p < 0.001). When all conduits were pooled, the overall effect estimate was neutral (0.08; 95% CI, –0.82 to 0.99; p = 0.86), although between-group differences were statistically significant (Q = 51.32; p = 0.00), reflecting heterogeneity in failure risks across conduit types.

**Figure 4.**
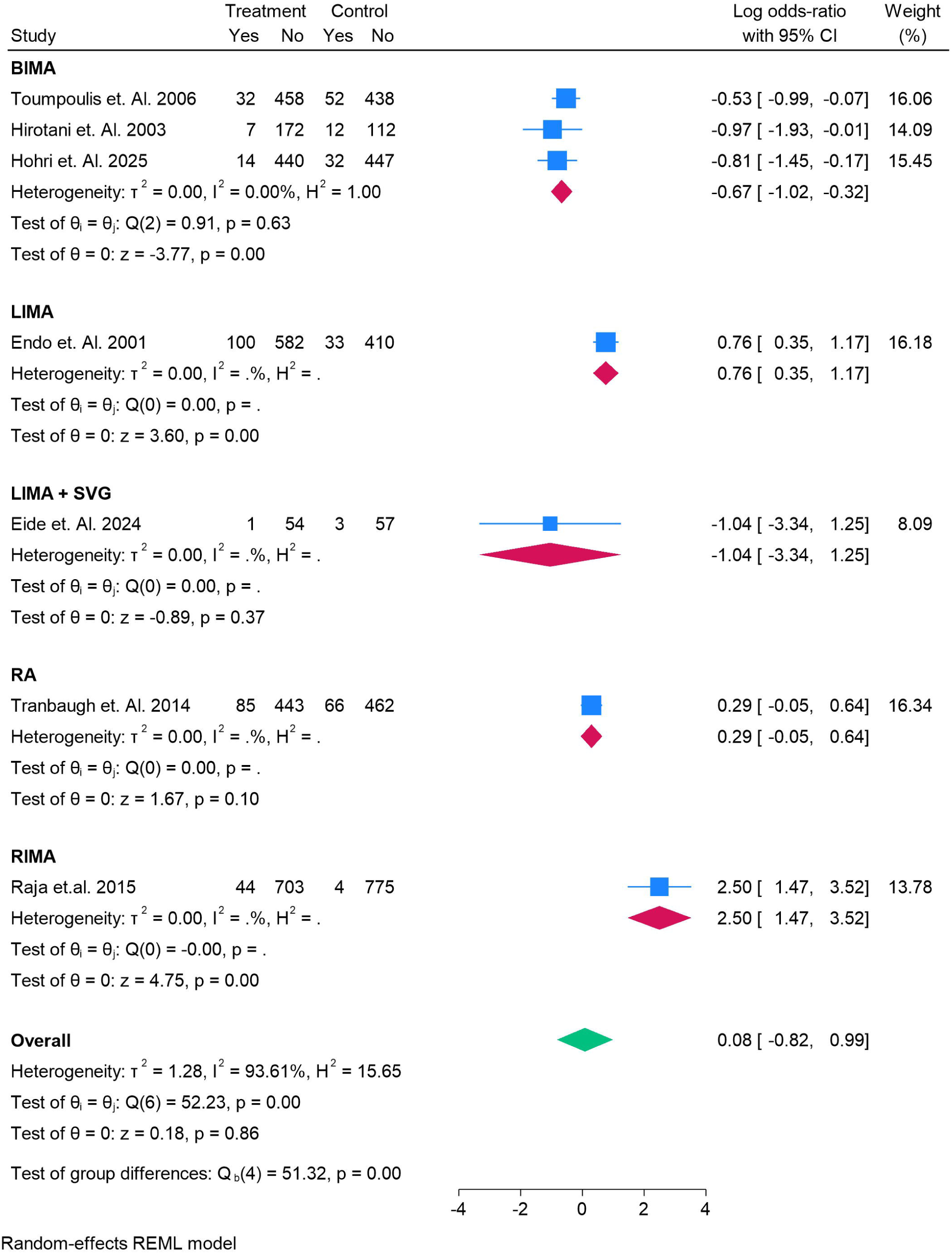
The odds of Grafts Failure of each coronary Graft in the CABG.

### Graft Patency Outcomes

Assessment of angiographic patency demonstrated variable results across conduit types (Figure 5). BIMA grafts showed a pooled mean difference of 6.50% (95% CI, –8.57 to 21.56; p = 0.40) compared with controls, though findings were highly heterogeneous (I² = 99.9%). LIMA grafts demonstrated wide variability across studies, with a pooled mean difference of –6.37% (95% CI, –28.37 to 15.63; p = 0.57), again with extreme heterogeneity (I² = 100%). By contrast, more consistent findings emerged with other conduits. LIMA + SVG grafting was associated with reduced patency (–1.00%; 95% CI, –1.54 to –0.46; p < 0.001). Similarly, radial artery (RA) grafts showed lower patency rates compared with controls (–4.00%; 95% CI, –4.44 to –3.56; p < 0.001), as did right internal mammary artery (RIMA) grafts (–4.00%; 95% CI, –4.13 to –3.87; p < 0.001). When all conduits were pooled, the overall estimate suggested no significant difference in patency compared with controls (–2.30%; 95% CI, –13.70 to 9.10; p = 0.69), but substantial heterogeneity was again evident (I² = 100%).

**Figure 5.**
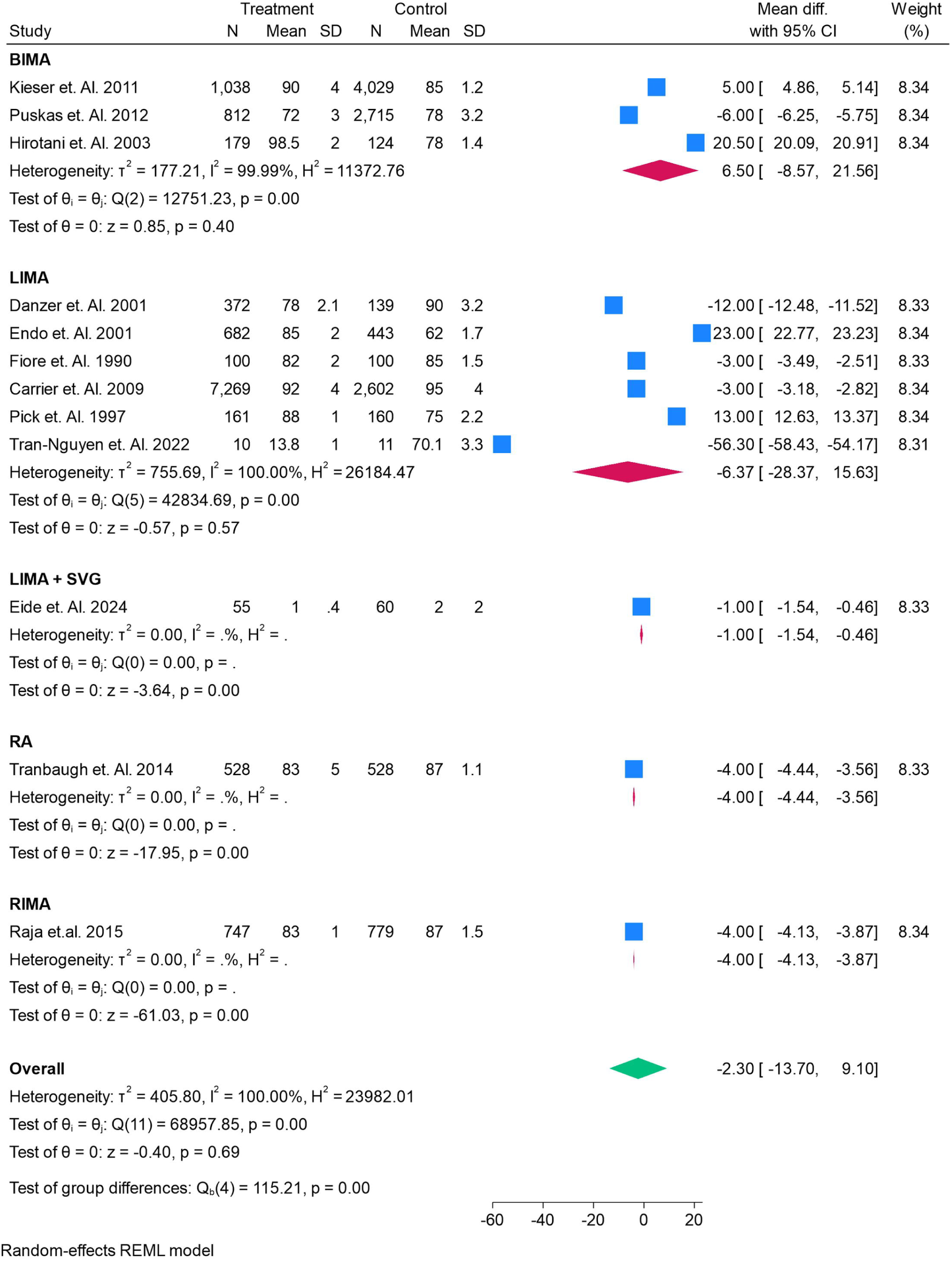
The percentage of each coronary Graft in the CABG for Patency of the Graft.

### Revascularization Outcomes

Analysis of repeat revascularization revealed mixed effects across conduit types (Figure 6). BIMA grafting demonstrated a non-significant reduction in the odds of revascularization, with a pooled log odds ratio of –0.35 (95% CI, –0.91 to 0.21; p = 0.22), although heterogeneity was moderate (I² = 83.1%). For LIMA grafts, the pooled effect estimate also did not reach significance (–0.51; 95% CI, –1.26 to 0.23; p = 0.18), though individual studies showed considerable variability, with some reporting reduced revascularization (e.g., Locker et al. 2012, OR –2.00; 95% CI, –2.77 to –1.23) and others showing neutral or adverse effects. The combination of LIMA with SVGs yielded no significant association (0.24; 95% CI, –0.79 to 1.27; p = 0.65). In contrast, RIMA grafting was associated with significantly higher odds of revascularization, with a pooled log odds ratio of 0.90 (95% CI, 0.51 to 1.30; p < 0.001). When all conduits were pooled, the overall estimate suggested no significant difference in the odds of repeat revascularization (–0.33; 95% CI, –0.81 to 0.15; p = 0.18), with substantial heterogeneity across studies (I² = 95.0%).

**Figure 6.**
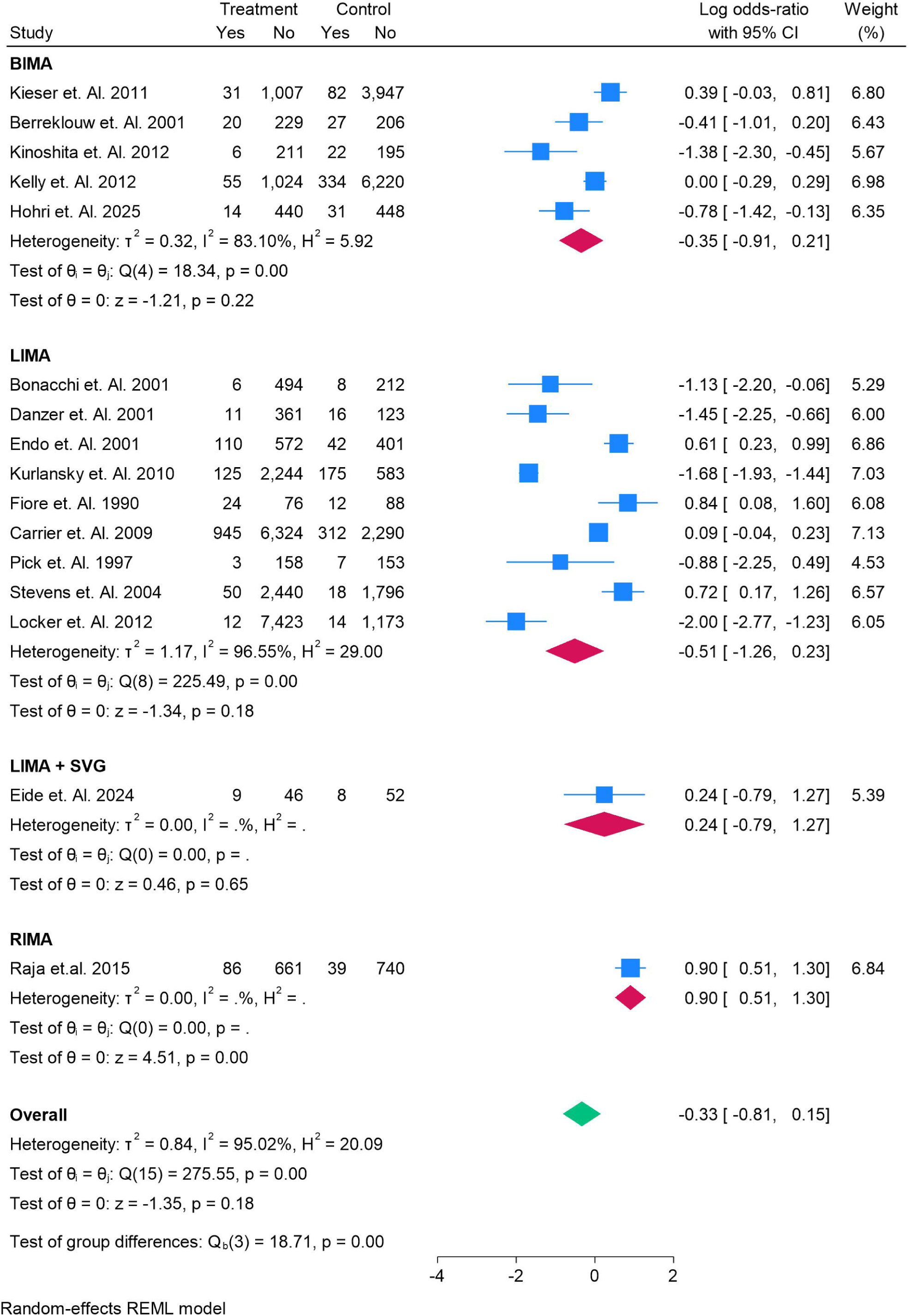
The odds of Revascularization of each coronary Graft in the CABG.

### All-Cause Mortality

Meta-analysis of all-cause mortality revealed distinct patterns across conduit types (Figure 7). BIMA grafting was associated with a significant survival advantage, with a pooled log odds ratio of –0.68 (95% CI, –0.93 to –0.44; p < 0.001), despite moderate heterogeneity (I² = 71.6%). In contrast, LIMA grafts alone were not significantly associated with reduced all-cause mortality, yielding a pooled effect estimate of 0.42 (95% CI, –0.32 to 1.17; p = 0.27), though individual studies demonstrated wide variability, ranging from protective to adverse associations. The combination of LIMA with SVGs did not significantly alter mortality risk (0.45; 95% CI, –0.20 to 1.10; p = 0.18). Radial artery (RA) grafts demonstrated a significant survival benefit, with a pooled log odds ratio of –0.84 (95% CI, –1.36 to –0.32; p = 0.002). In contrast, RIMA grafting was associated with increased mortality, with a pooled estimate of 1.09 (95% CI, 0.60 to 1.58; p < 0.001). When all conduits were pooled, the overall effect estimate was neutral (–0.10; 95% CI, –0.49 to 0.28; p = 0.60), with substantial heterogeneity across studies (I² = 94.9%).

**Figure 7.**
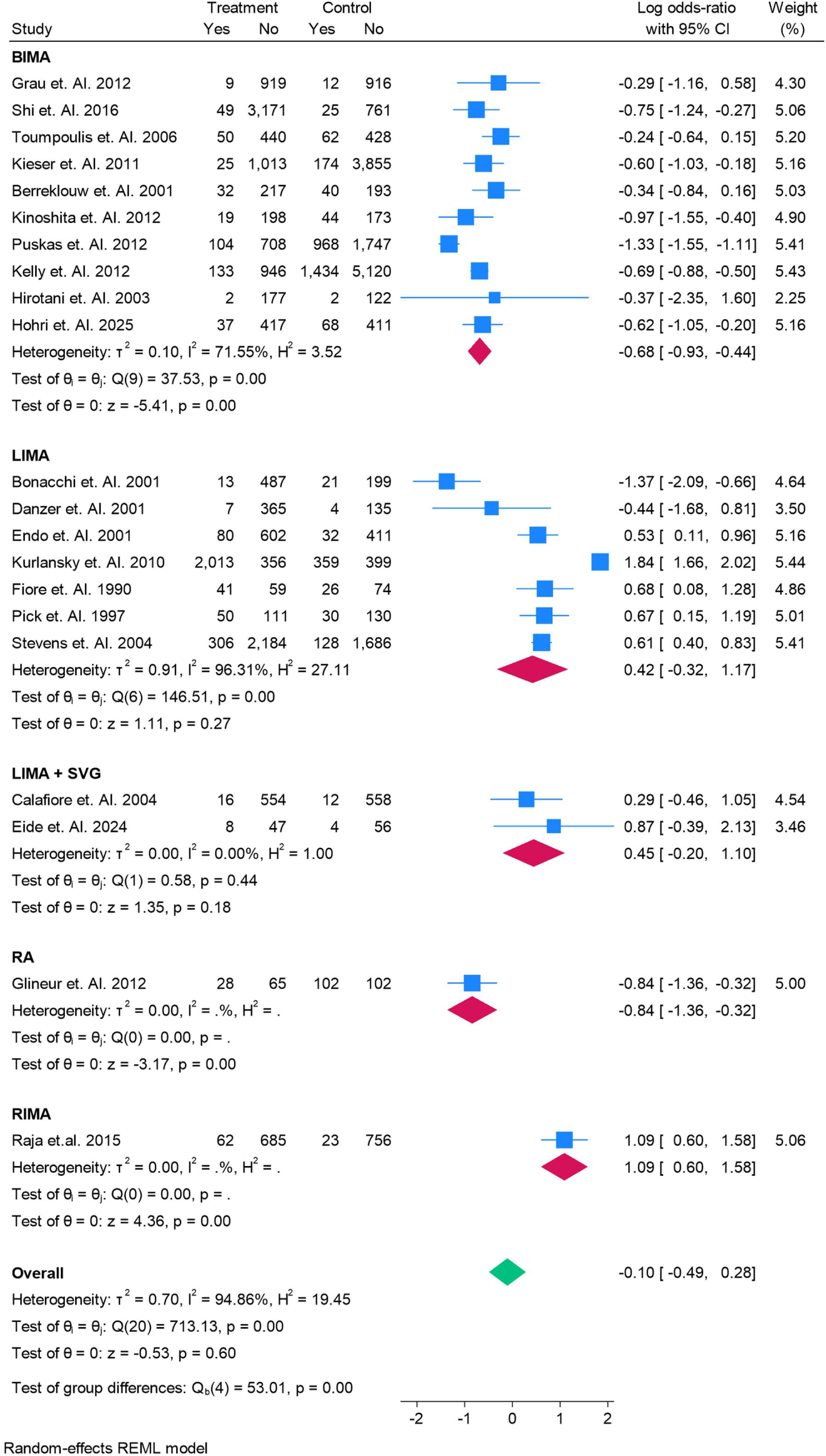
The odds of All Cause Mortality of each coronary Graft in the CABG.

### Cardiac Mortality

The pooled analysis of cardiac mortality demonstrated variable outcomes across conduit types (Figure 8). BIMA grafting was associated with a non-significant reduction in cardiac mortality, with a pooled log odds ratio of –0.84 (95% CI, –1.83 to 0.15; p = 0.10), with moderate heterogeneity (I² = 55.9%). For LIMA grafts, no significant association was observed overall (0.15; 95% CI, –0.69 to 0.99; p = 0.73), though individual studies showed heterogeneity, ranging from protective effects (Bonacchi et al., log OR –1.35) to increased risk (Mohammadi et al., log OR 0.76). The combination of LIMA with SVGs yielded no significant effect on cardiac mortality (0.63; 95% CI, –0.30 to 1.56; p = 0.18). In contrast, radial artery (RA) grafts demonstrated a significant reduction in cardiac mortality, with a pooled log odds ratio of –1.30 (95% CI, –1.89 to –0.71; p < 0.001).When all conduit types were pooled, the overall estimate was neutral (–0.20; 95% CI, –0.85 to 0.45; p = 0.55), with considerable heterogeneity (I² = 87.2%).

**Figure 8.**
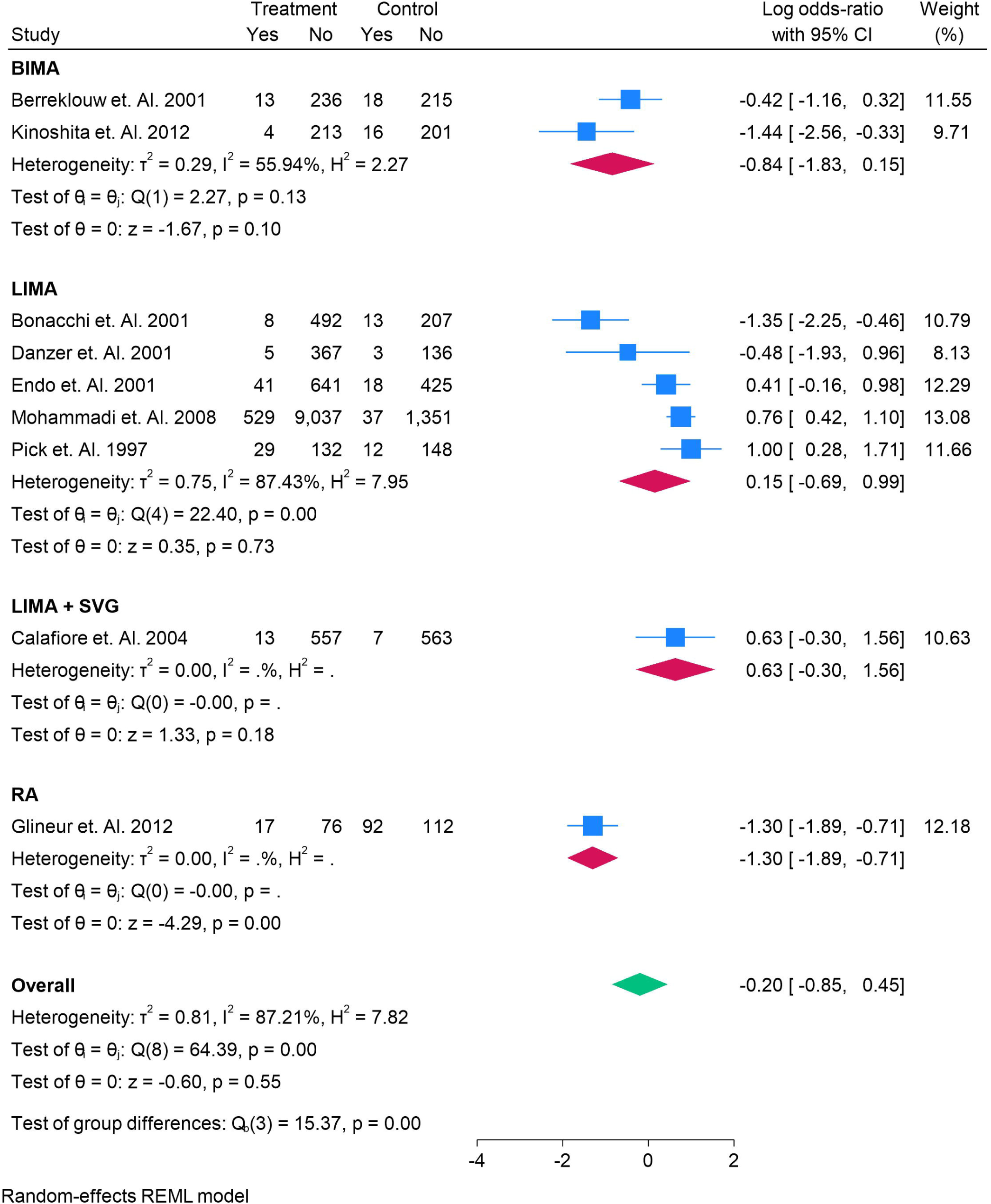
The odds of Cardiac Mortality of each coronary Graft in the CABG.

### Adverse Events

Analysis of overall adverse events demonstrated mixed findings across graft types (Figure 9). BIMA grafting was associated with a non-significant reduction in adverse events, with a pooled log odds ratio of – 0.32 (95% CI, –0.83 to 0.19; p = 0.22), though moderate heterogeneity was observed (I² = 90.2%). LIMA grafts also showed no significant effect (–0.12; 95% CI, –1.30 to 1.06; p = 0.84), with substantial variability between studies. The combination of LIMA + SVGs similarly showed no association with adverse event risk (0.11; 95% CI, –0.40 to 0.62; p = 0.67). By contrast, radial artery (RA) grafts were associated with a significant reduction in adverse events, with a pooled log odds ratio of –0.69 (95% CI, –1.09 to –0.28; p < 0.01). When all grafts were analyzed together, the overall pooled effect suggested no statistically significant difference in adverse event risk (–0.25; 95% CI, –0.65 to 0.15; p = 0.23), with high heterogeneity across studies (I² = 92.2%).

**Figure 9.**
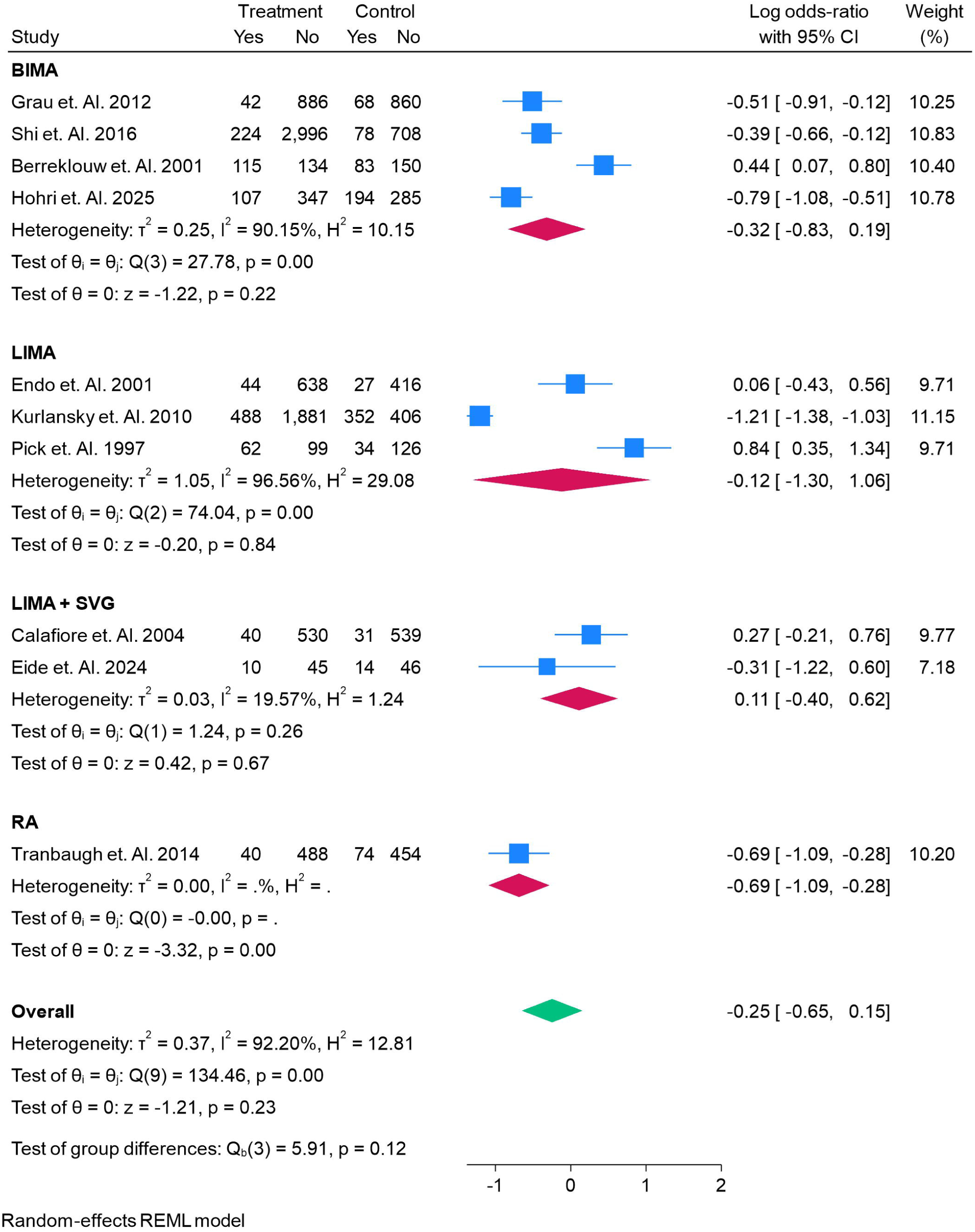
The odds of All Adverse Events of each coronary Graft in the CABG.

### Sternal Wound Infections

The analysis of sternal wound infections demonstrated important variation by conduit type (Figure 10). BIMA grafting was associated with a non-significant increase in the odds of sternal wound infection (log OR 0.13; 95% CI, –0.53 to 0.78; p = 0.70), with moderate heterogeneity across studies (I² = 72.5%). LIMA grafts showed no significant association with infection (–0.07; 95% CI, –0.64 to 0.50; p = 0.81), with low to moderate heterogeneity (I² = 61.2%). The combination of LIMA with SVGs similarly demonstrated no significant effect (–0.66; 95% CI, –2.09 to 0.78; p = 0.37). By contrast, radial artery (RA) grafts were associated with a directionally protective effect, with a pooled estimate of –1.75 (95% CI, –3.67 to 0.18; p = 0.08), though this did not reach statistical significance. RIMA grafting did not significantly influence infection risk (0.25; 95% CI, –0.40 to 0.89; p = 0.46). Notably, SVGs were associated with a significant reduction in wound infection, with a pooled log odds ratio of –1.77 (95% CI, –2.61 to –0.92; p < 0.001). The overall pooled estimate across all conduits indicated no significant difference in sternal wound infection risk (–0.22; 95% CI, –0.64 to 0.20; p = 0.31), though between-group differences were statistically significant (Q = 18.98; p < 0.001).

**Figure 10.**
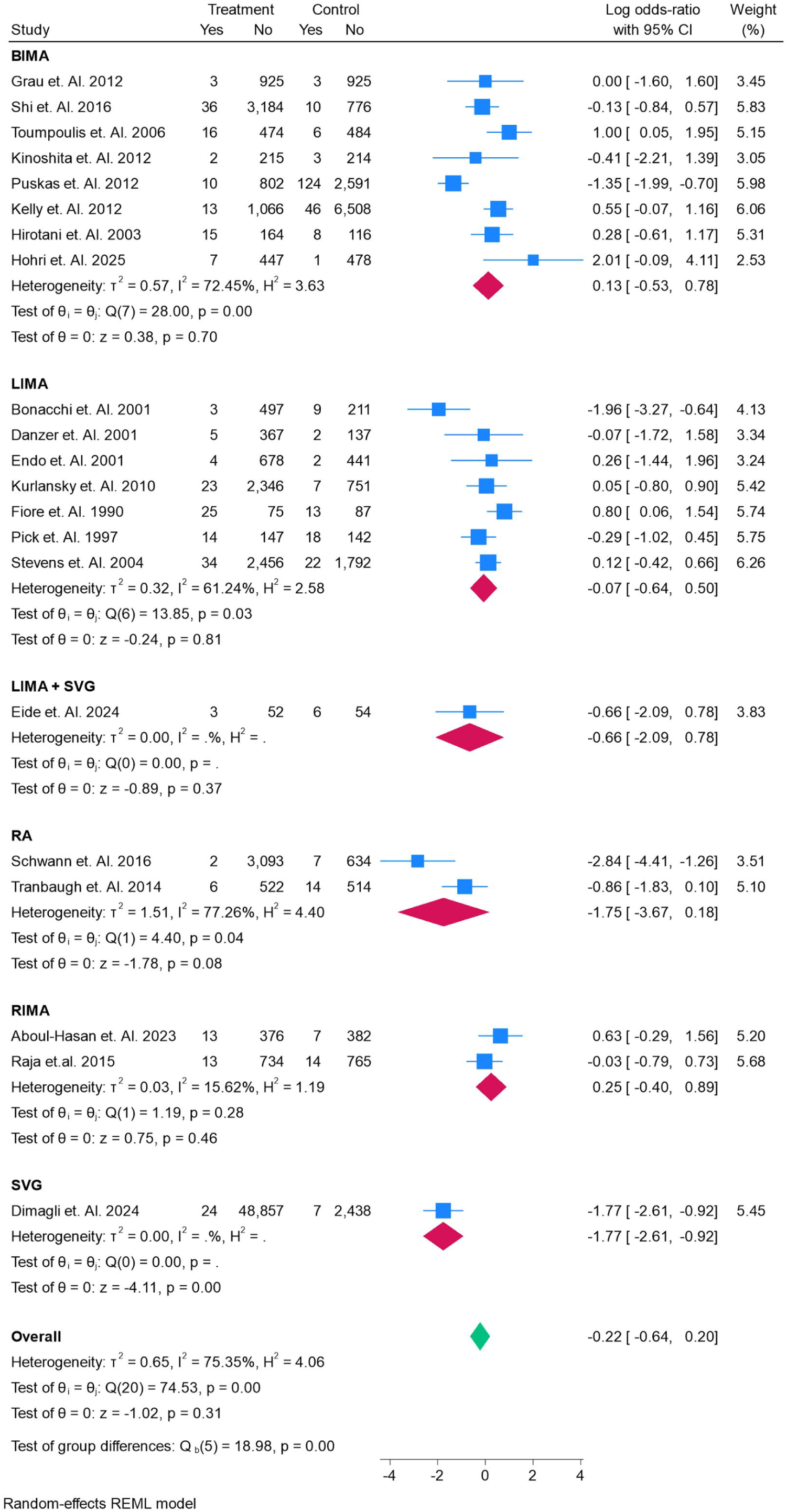
The odds of Sternal Wound Infections of each coronary Graft in the CABG.

### Length of Stay

Analysis of postoperative length of stay revealed variability across graft types (Figure 11). BIMA grafting was associated with a modest but statistically significant reduction in hospital stay, with a pooled mean difference of –0.83 days (95% CI, –1.45 to –0.20; p = 0.01). In contrast, LIMA grafts demonstrated no significant association, with a pooled estimate of –0.31 days (95% CI, –1.86 to 1.24; p = 0.70), though heterogeneity was extreme (I^²^ = 99.7%). The combination of LIMA with SVGs was associated with an increased length of stay (+2.20 days; 95% CI, –2.48 to 6.88; p = 0.36), though estimates were imprecise due to limited data. For SVGs, no difference in length of stay was observed (0.00 days; 95% CI, –0.05 to 0.05; p = 1.00). When all grafts were analyzed collectively, the overall pooled estimate was neutral (–0.49 days; 95% CI, –1.11 to 0.13; p = 0.12), with substantial heterogeneity across studies (I^²^ = 99.3%).

**Figure 11.**
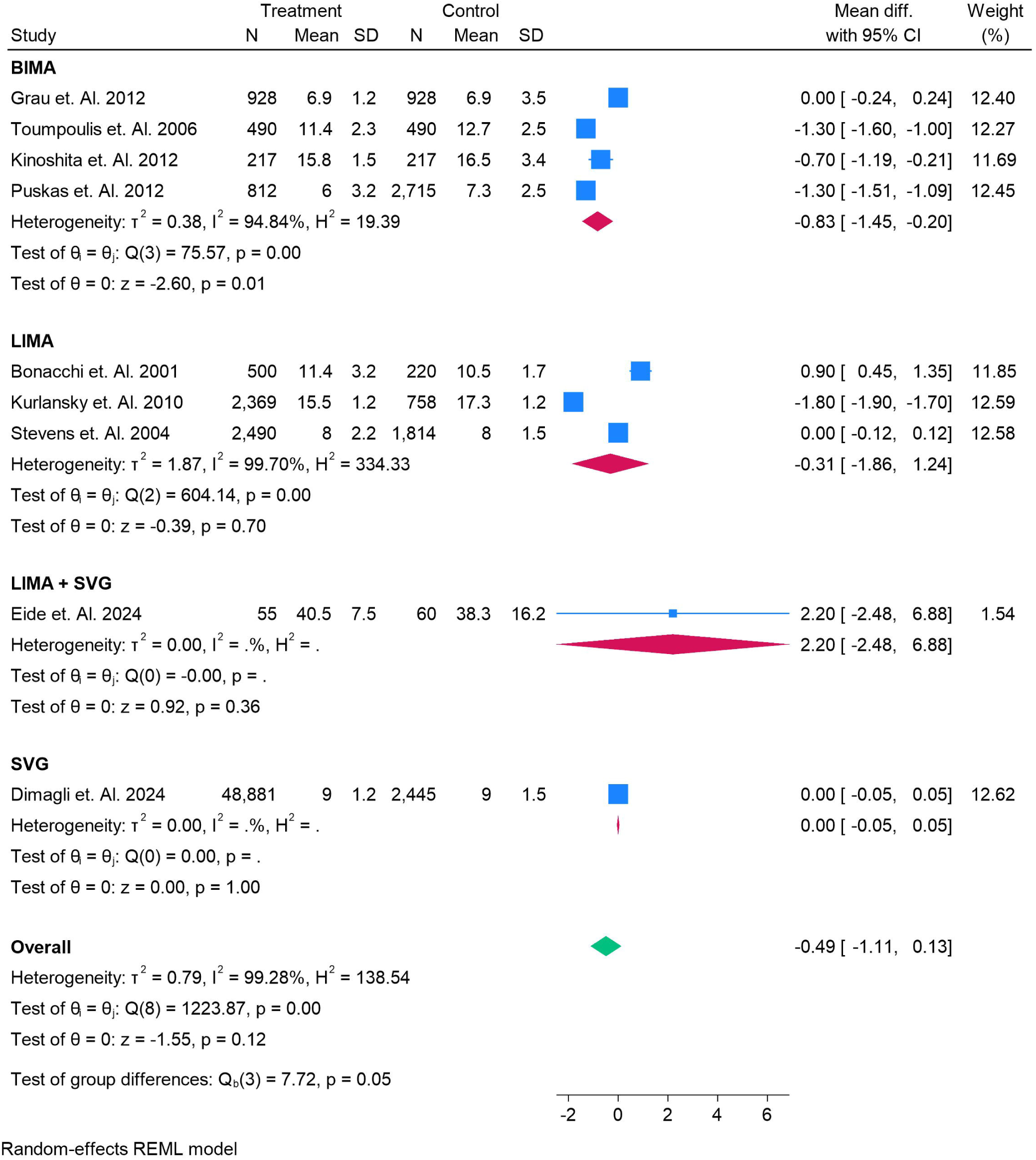
The Mean Difference of each coronary Graft 1n the CABG for Length of Stay.

### Worsening of Renal Function

Analysis of postoperative renal outcomes revealed no significant differences between graft types (Figure 12). BIMA grafting was not associated with a change in renal function, with a pooled mean difference of –3.06 (95% CI, –39.46 to 33.34; p = 0.87), and no evidence of heterogeneity (I² = 0%). Similarly, LIMA grafts demonstrated no significant association, with a pooled mean difference of 5.16 (95% CI, –38.15 to 48.47; p = 0.82). The combination of LIMA with SVGs also showed no effect (0.00; 95% CI, –20.73 to 20.73; p = 1.00). For radial artery (RA) grafts, the pooled estimate was neutral (0.00; 95% CI, –63.33 to 63.33; p = 1.00). RIMA grafts similarly showed no significant effect (2.68; 95% CI, –41.09 to 46.46; p = 0.90). The single large study reporting outcomes for SVGs yielded an imprecise estimate with wide confidence intervals (88.00; 95% CI, –1844.99 to 2020.99; p = 0.93). When all conduits were pooled, the overall estimate was neutral (0.43; 95% CI, –14.67 to 15.53; p = 0.96), with no evidence of heterogeneity (I² = 0%).

**Figure 12.**
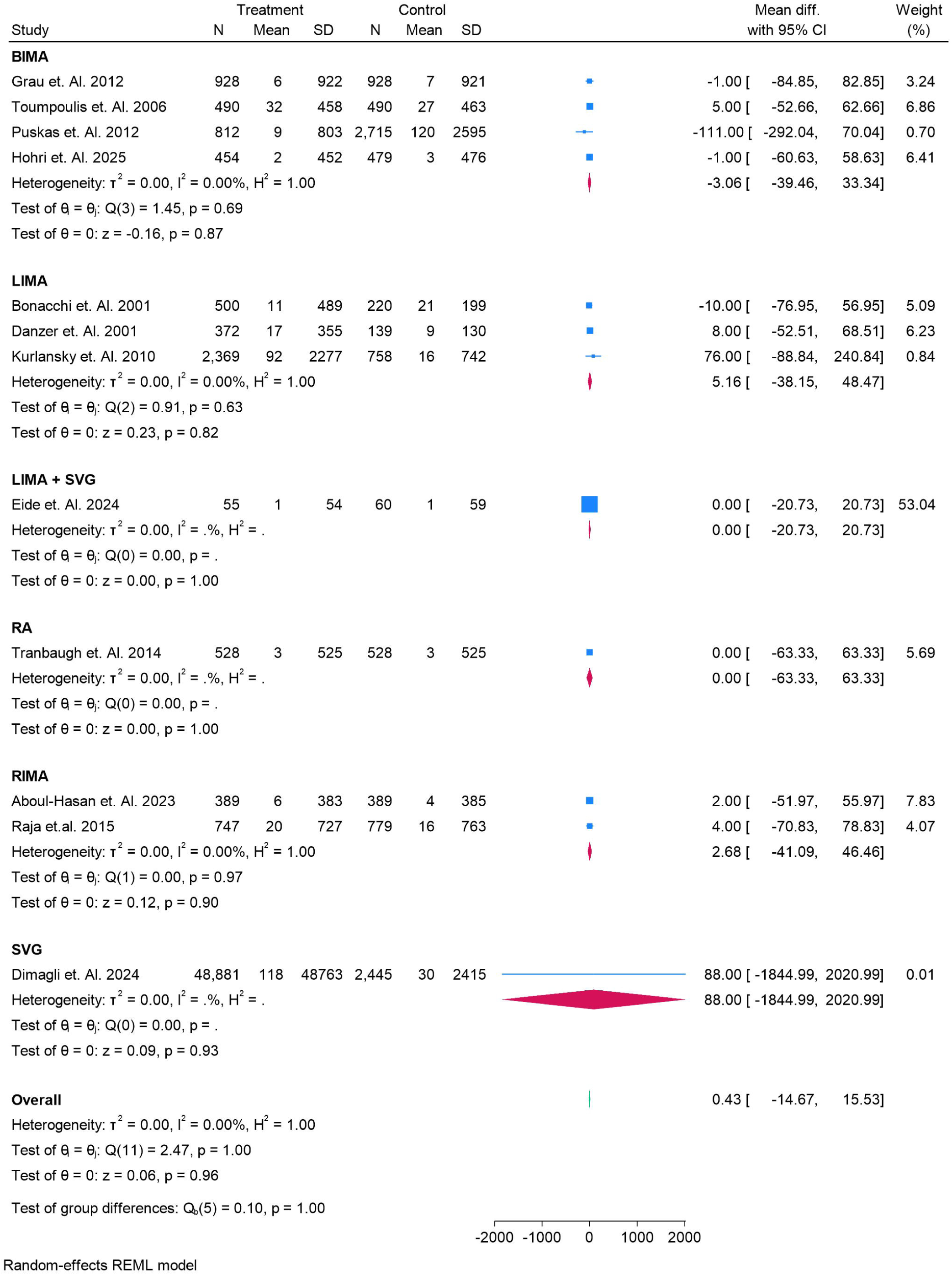
The odds of Worsening of Renal Function Ill each coronary Graft in the CABG.

## Discussion

This comprehensive meta-analysis of 32 observational studies, encompassing more than 170,000 patients, provides an integrated assessment of the comparative performance of different coronary artery bypass grafts (CABG). The findings highlight the nuanced balance between survival benefits, graft patency, complication risks, and long-term outcomes across various conduits.

Our results confirm the consistent superiority of bilateral internal mammary artery (BIMA) grafting, which was associated with improved survival and reduced graft failure. These findings align with prior evidence demonstrating that multiple arterial grafts confer a survival advantage compared with single internal thoracic artery grafting [54]. Although the Arterial Revascularization Trial (ART) did not show a clear survival difference between BIMA and SITA at 10 years [55], subsequent meta-analyses and large observational studies have reinforced the durability and long-term benefits of BIMA [56]. However, increased perioperative risk of sternal wound infection, particularly in diabetics and obese patients, has limited its widespread adoption [57]. Our analysis also observed trends toward higher wound complications in BIMA, underscoring the importance of careful patient selection.

The left internal mammary artery (LIMA) to the left anterior descending artery remains the cornerstone of CABG, with well-documented benefits in long-term patency and survival [58]. While our pooled estimates did not show consistent survival superiority for LIMA alone, this likely reflects its near-universal use as the default graft in most patients, rendering comparative analyses less meaningful. The combination of LIMA with saphenous vein grafts (SVGs) remains the most common strategy worldwide, offering technical feasibility and adequate revascularization, though at the cost of reduced long-term durability.

Radial artery (RA) grafts were associated with reduced cardiac mortality and lower adverse events in our study, corroborating previous evidence that RA offers superior patency compared with SVGs, especially when used to bypass high-grade stenoses [59]. Recent individual patient data meta-analyses have also confirmed improved long-term outcomes with RA over venous conduits [60]. These findings support increasing adoption of RA grafts, provided that adequate patient selection and antispasm protocols are applied.

Conversely, saphenous vein grafts were consistently associated with worse survival and higher long-term failure, confirming their vulnerability to atherosclerosis and occlusion [61]. While technically simple and versatile, their inferior durability highlights the pressing need for optimizing medical therapy and vein harvesting techniques to improve outcomes when SVGs are used.

Interestingly, right internal mammary artery (RIMA) results were mixed, with some studies reporting increased mortality or revascularization risk. This contrasts with prior analyses suggesting comparable patency to LIMA when used in situ [62]. The heterogeneity likely reflects differences in patient populations, surgical expertise, and grafting strategies.

Several limitations must be acknowledged. First, all included studies were observational, and residual confounding cannot be excluded despite adjustments. Second, heterogeneity across studies was substantial for several outcomes, reflecting differences in populations, follow-up durations, and surgical techniques. Third, reporting of secondary outcomes such as renal dysfunction, adverse events, and length of stay was limited and inconsistent.

Despite these limitations, this meta-analysis provides robust evidence that arterial grafts—particularly BIMA and RA—offer superior long-term outcomes compared with SVGs. The findings support a shift toward greater use of multiple arterial conduits in CABG, while emphasizing the importance of individualized decision-making based on patient comorbidities, anatomy, and surgical expertise. Future randomized controlled trials with long-term follow-up are needed to resolve ongoing uncertainties, particularly regarding the role of RIMA and total arterial revascularization strategies.

## Conclusion

This meta-analysis of over 170,000 patients demonstrates that arterial grafts, particularly BIMA and the radial artery, provide superior long-term outcomes in CABG compared with saphenous vein grafts. While LIMA remains the cornerstone conduit, expanding the use of multiple arterial grafting may further enhance survival and durability. Nonetheless, risks such as sternal wound complications with BIMA and variable outcomes with RIMA necessitate individualized decision-making. These findings underscore the importance of optimizing graft selection to balance efficacy and safety, while highlighting the need for future randomized trials to clarify the role of total arterial revascularization strategies.

## Conflict of Interest

The authors certify that there is no conflict of interest with any financial organization regarding the material discussed in the manuscript.

## Funding

The authors report no involvement in the research by the sponsor that could have influenced the outcome of this work.

## Authors’ contributions

All authors contributed equally to the manuscript and read and approved the final version of the manuscript.

## Data Availability

Supplementary file

## Notes

### Competing Interest Statement

The authors have declared no competing interest.

### Clinical Protocols

https://www.crd.york.ac.uk/PROSPERO/view/CRD420251151159

