## supplementary file for "Comparative Efficacy and Safety of Coronary Artery Bypass Grafts: Insights from a Systematic Review and Meta-Analysis"

Supplementary Files

Search String

(("Coronary Artery Bypass"[Mesh] OR CABG[tiab] OR "coronary artery bypass grafting"[tiab])

AND

(("Right Internal Mammary Artery"[tiab] OR RIMA[tiab] OR "Internal Mammary Arteries"[Mesh] OR "Mammary Artery"[tiab])

OR ("Radial Artery"[Mesh] OR "Radial Artery"[tiab] OR RA[tiab])

OR ("Saphenous Vein"[Mesh] OR "Saphenous Vein Graft"[tiab] OR SVG[tiab])

OR ("Gastroepiploic Artery"[Mesh] OR "Gastroepiploic Artery"[tiab] OR GEA[tiab])

OR ("Inferior Epigastric Artery"[tiab] OR IEA[tiab]))

AND

(conduit[tiab] OR graft[tiab] OR bypass[tiab] OR revascularization[tiab] OR reconstruction[tiab]))

Table 1. Summary Findings of the given studies

| **References** | **Author Year** | **Country** | **Type of Study** | **Total** | **Male** | **Female** | **Follow Up** | **On vs Off Pump** | **Graft** | **Graft number** | **Graft 2** | **Graft 2 Number** | **GRADE** | **Main Finding** |
| --- | --- | --- | --- | --- | --- | --- | --- | --- | --- | --- | --- | --- | --- | --- |
| 22 | Bonacchi et. Al. 2001 | Italy | Retrospective Observational | 576 | 457 | 119 | 32.4 | On-Pump | LIMA | 500 | BIMA | 220 | High | Use of bilateral internal mammary arteries in urgent CABG for unstable angina is feasible and associated with improved long-term survival without increased operative risk compared to single IMA grafting |
| 23 | Calafiore et. Al. 2004 | Italy | propensity score–matched cohort analysis | 1602 | 930 | 210 | 87.6 | On-Pump | LIMA + SVG | 570 | BIMA + SVG | 570 | High | Bilateral internal mammary artery (BIMA) grafting provided significantly better 10-year freedom from cardiac death, myocardial infarction, and target cardiac events compared with single LIMA plus saphenous vein grafts. |
| 24 | Danzer et. Al. 2001 | Switzerland | Retrospective observational cohort study | 694 | 619 | 75 | 120 | On-Pump | ITA | 372 | ITA + SVG | 139 | Moderate | Bilateral internal thoracic artery grafting significantly reduced cardiac mortality and reintervention rates without increasing postoperative complications compared with single ITA grafting |
| 25 | Endo et. Al. 2001 | Japan | Retrospective observational cohort study | 1131 | 556 | 956 | 175 | On-Pump | SIMA | 682 | BIMA | 443 | Moderate | BIMA grafting showed significantly higher graft patency (97.3% vs 94.3%) and lower need for revascularisation compared with SIMA, with similar mortality and complication rates |
| 26 | Grau et. Al. 2012 | USA | Retrospective, propensity-matched cohort study | 1856 | 828 | 829 | 108 | Off-Pump | BIMA | 928 | LIMA + SVG | 928 | Moderate | BIMA grafting confers significantly improved long-term survival compared to LIMA-SVG, especially when performed off-pump, without increasing perioperative complications |
| 27 | Glineur et. Al. 2012 | Belgium | Porspective Cohort | 588 | 506 | 82 | 193 | Off-Pump | BITA + RGEA | 93 | BITA + SVG | 204 | Moderate | Use of a third arterial graft (RGEA) with BITA significantly improves long-term survival and cardiac survival compared to SVG |
| 28 | Pettinari et. Al. 2015 | Belgium | Retrospective observational cohort study | 3496 | 1748 | 1748 | 120 | On-Pump | BITA | 1328 | SITA | 2168 | Moderate | Bilateral internal thoracic artery (BITA) grafting increases long-term (10-year) survival compared with single internal thoracic artery (SITA) grafting in elderly patients (>70 years) |
| 29 | Shi et. Al. 2016 | Australia | Retrospective, propensity-matched cohort study | 4006 | 3080 | 926 | 180 | On-Pump | RA + ITA | 3220 | SV + ITA | 786 | High | Bilateral internal thoracic artery (BITA) grafting increases long-term (10-year) survival compared with single internal thoracic artery (SITA) grafting in elderly patients (>70 years) |
| 30 | Schwann et. Al. 2016 | Multicenter | Multi-institutional, retrospective observational study | 8220 | 5851 | 2369 | 189 | On-Pump | RA | 3095 | RITA | 641 | High | Both **radial artery (RA)** and **right internal thoracic artery (RITA)** used as second conduits with LITA provide **equivalent long-term survival (~16 years) and superior outcomes compared to single LITA grafts**, though RITA increases sternal wound infection risk |
| 31 | Toumpoulis et. Al. 2006 | USA | Retrospective, propensity-matched cohort study | 980 | 545 | 435 | 56.4 | On-Pump | BITA | 490 | SITA | 490 | Moderate | BITA grafting did not improve overall long-term survival compared with SITA in diabetics, but patients aged 60–69 benefited, whereas those >79 years did worse with BITA |
| 32 | Kurlansky et. Al. 2010 | USA | Retrospective, propensity-matched cohort study | 4584 | 3602 | 982 | 133 | On-Pump | SIMA | 2369 | BIMA | 758 | Moderate | Bilateral internal mammary artery (BIMA) grafting significantly improves long-term survival compared with single internal mammary artery (SIMA), with lower morbidity and acceptable perioperative risk |
| 33 | Kieser et. Al. 2011 | Canada | Observational Cohort Study | 5601 | 2800 | 2800 | 94.8 | On-Pump | BITA | 1038 | SITA | 4029 | Moderate | BITA grafting was associated with lower crude mortality and revascularisation compared to SITA, with benefits most evident in patients younger than 70 years |
| 34 | Mohammadi et. Al. 2008 | Canada | Retrospective observational cohort study | 12231 | 7180 | 5051 | 68.4 | On-Pump | SITA | 9566 | BITA | 1388 | Moderate | The survival benefit of bilateral ITA over single ITA grafting is significant only up to age 60; after that, the additional benefit is lost, though a single ITA remains superior to vein grafts in all age groups |
| 35 | Fiore et. Al. 1990 | USA | Retrospective comparative study | 200 | 100 | 100 | 172.8 | On-Pump | SITA | 100 | BITA | 100 | Moderate | Double ITA grafting showed significantly higher long-term survival, patency, and freedom from ischemic events compared to single ITA grafting |
| 36 | Berreklouw et. Al. 2001 | Netherlands | Retrospective, matched cohort study | 482 | 418 | 64 | 118 | On-Pump | BITA | 249 | SITA | 233 | Moderate | Bilateral internal thoracic artery grafting significantly improves ischemic event-free survival compared with single LITA grafting, though overall survival is similar at 13 years |
| 37 | Lytle et. Al. 2004 | USA | Retrospective, propensity-matched cohort study | 10124 | 8706 | 1418 | 194 | On-Pump | BITA | 1152 | SITA | 1152 | Moderate | Bilateral internal thoracic artery grafting significantly improves 20-year survival compared with single internal thoracic artery grafting (50% vs 37%, p<0.0001). |
| 38 | Carrier et. Al. 2009 | Canada | Retrospective observational cohort study | 6655 | 4877 | 1778 | 60 | On-Pump | LIMA | 7269 | BITA | 2602 | Moderate | Early statin therapy after CABG improves long-term survival in single ITA patients, equalizing outcomes with bilateral ITA grafts |
| 39 | Kinoshita et. Al. 2012 | Japan | Retrospective, propensity-matched cohort study | 491 | 336 | 98 | 51.6 | Off-Pump | BITA | 217 | SITA | 217 | High | In elderly CABG patients, **off-pump bilateral ITA grafting significantly improved survival and reduced cardiac events compared with single ITA, without increasing operative risk** |
| 40 | Puskas et. Al. 2012 | USA | Retrospective, matched cohort study | 3527 | 2501 | 1026 | 96 | Off-Pump | BITA | 812 | SITA | 2715 | High | Bilateral internal thoracic artery grafting (BITA) significantly improves long-term survival compared to single internal thoracic artery grafting (SITA), without increasing major adverse events, even in diabetic patients. |
| 41 | Dimagli et. Al. 2024 | UK | Retrospective observational cohort study | 58063 | 0 | 58063 | 4 | On-Pump | SVG | 48881 | RA | 2445 | Moderate | **In women undergoing CABG, the radial artery had outcomes comparable to saphenous vein grafts, while the right internal thoracic artery was associated with higher in-hospital mortality and more sternal wound infections.** |
| 42 | Pick et. Al. 1997 | USA | Retrospective observational cohort study | 481 | 161 | 320 | 118 | On-Pump | SIMA | 161 | BIMA | 160 | High | Bilateral internal thoracic artery (BITA) plus vein grafting offers superior long-term survival and event-free outcomes compared with single ITA (SITA) plus vein grafting in multivessel CABG. |
| 43 | Stevens et. Al. 2004 | Canada | Retrospective observational cohort study | 4382 | 3471 | 835 | 132 | On-Pump | SITA | 2490 | BITA | 1814 | Moderate | Bilateral internal thoracic artery (BITA) plus vein grafting offers superior long-term survival and event-free outcomes compared with single ITA (SITA) plus vein grafting in multivessel CABG. |
| 44 | Kelly et. Al. 2012 | Canada | Retrospective observational cohort study | 8264 | 6248 | 2019 | 56.4 | Off-Pump | BITA | 1079 | SITA | 6554 | Moderate | Bilateral internal thoracic artery (BITA) grafting offers a significant long-term survival advantage over single internal thoracic artery (SITA) grafting in CABG patients |
| 45 | Parsa et. Al. 2013 | USA | Retrospective observational cohort study | 19482 | 3897 | 15585 | 300 | On-Pump | SITA | 16881 | BITA | 728 | Moderate | *Over 25 years, using multiple internal thoracic arteries (MITA) reduced major adverse cardiac events by ~30% vs no ITA and by ~11% vs SITA, but mortality difference between SITA and MITA was minimal.* |
| 46 | Tranbaugh et. Al. 2014 | USA | Retrospective, propensity-matched cohort study | 2488 | 1940 | 548 | 108 | On-Pump | RA | 528 | RITA | 528 | Moderate | Radial artery (RA) grafting has similar long-term survival and patency to right internal thoracic artery (RITA) grafting but is associated with fewer major adverse events, especially in high-risk patients (diabetes, COPD, obesity, older age). |
| 47 | Aboul-Hasan et. Al. 2023 | Poland | Retrospective, propensity-matched cohort study | 1198 | 618 | 160 | 90.4 | On-Pump | RITA | 389 | RA | 389 | Moderate | The use of **RITA or RA as the second arterial conduit during CABG yields comparable long-term survival and MACCE outcomes**, making the choice dependent on patient characteristics and surgeon preference |
| 48 | Raja et.al. 2015 | UK | Retrospective, propensity-matched cohort study | 1526 | 779 | 747 | 96 | On-Pump | RIMA | 747 | RA | 779 | Moderate | RIMA as a second conduit improved long-term survival and reduced repeat revascularization compared to RA, without increasing perioperative complications |
| 49 | Hirotani et. Al. 2003 | Japan | Retrospective comparative study | 303 | 230 | 73 | 1 | On-Pump | BITA | 179 | SITA | 124 | Moderate | In diabetic patients undergoing CABG, **bilateral ITA grafting was not associated with higher morbidity or mortality compared to unilateral ITA, but also did not show a significant survival or event-free advantage** |
| 50 | Eide et. Al. 2024 | Germany | Retrospective observational cohort study | 115 | 88 | 27 | 60.3 | On-Pump | LIMA + SVG | 55 | LIMA + RA | 60 | Moderate | CABG with a composite T-graft between LIMA and saphenous vein showed **comparable medium-term morbidity and mortality outcomes** to LIMA + radial artery grafting, with no significant difference in survival or adverse events |
| 51 | Tran-Nguyen et. Al. 2022 | Canada | Prospetive Observational Study | 10 | 9 | 1 | 1 | Off-Pump | LIMA | 10 | SVG | 11 | Moderate | **Arterial grafts, especially LIMA, showed significantly lower abnormal wall shear stress compared to venous grafts (SVG), suggesting hemodynamic mechanisms may underlie their superior long-term patency.** |
| 52 | Locker et. Al. 2012 | USA | Retrospective observational cohort study | 8622 | 5594 | 1008 | 91 | On-Pump | LIMA | 7435 | RA | 1187 | Low | Use of multiple arterial grafts (MultArt) in CABG significantly improves 15-year survival compared with conventional LIMA + saphenous vein grafting |
| 53 | Hohri et. Al. 2025 | USA | Retrospective observational cohort study | 933 | 727 | 206 | 55.6 | Off-Pump | BIMA | 454 | LIMA | 479 | Low | BIMA grafting in patients with mildly decreased renal function undergoing CABG was associated with better 6-year survival and lower major adverse cardiac or cerebrovascular events compared to SIMA grafting, but with higher risk of sternal wound infection |

Risk of Bias

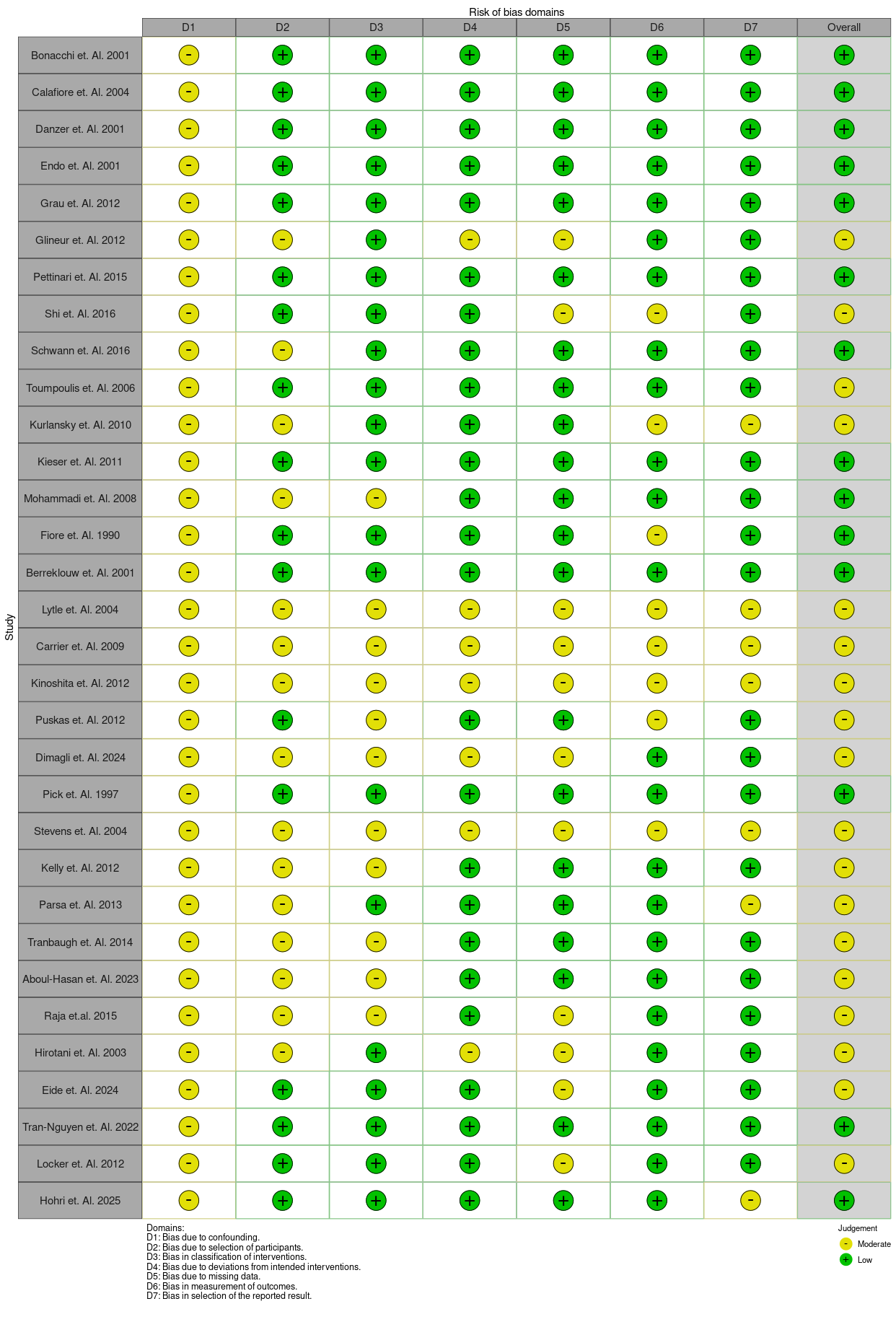
